# Hospital pharmacy leader perspectives on advocating for clinical pharmacy services: A national survey

**DOI:** 10.64898/2026.01.26.26344866

**Authors:** Huong Duong, Morgan Karnbach, Chelsea Keedy, Kelli Henry, Mojdeh Heavner, Brian Murray, Mary Ghaffari, Jennifer Majchrzak, Meghan D. Swarthout, Andrea Sikora, Susan Smith

## Abstract

**Purpose:** Although numerous research studies have demonstrated the positive impact of clinical pharmacy services, these benefits do not translate into sustained practice changes without support from hospital pharmacy leaders. Factors influencing leadership decisions to expand pharmacy services remained unclear. This study aimed to identify barriers to implementing pharmacy practice model changes and gain insights on potential methods of overcoming these barriers from the hospital pharmacy leader perspective.

**Summary:** We conducted a national, cross-sectional survey of hospital pharmacy leaders using the REDCap platform, distributed via email over three weeks between September to October 2025. The survey included questions about perceptions related to implementation of practice model changes, resources/evidence used to justify clinical positions, barriers to expanding clinical pharmacy services, and demographics of healthcare systems they represented. The survey included Likert-scales and open-ended questions. The primary outcome was types of evidence most compelling to justify clinical pharmacist positions. Secondary outcomes included resources currently in use for decision-making and perceived barriers. The survey highlighted key factors influencing administrative decision-making regarding the expansion of clinical pharmacy services and revealed significant barriers to justifying clinical positions related to knowledge gaps, underscoring the need for further research to develop evidence-based metrics that capture the comprehensive benefits that clinical pharmacists can offer.

**Conclusion:** This survey provided valuable insights into hospital pharmacy leader perspectives on resources and evidence needed to support expanded pharmacy services and justify clinical pharmacist positions. These insights can inform future research by ensuring that metrics that are both clinically and administratively significant are included in outcomes.

**Key points:**

1. While clinical pharmacy services are known to improve patient care, their benefits do not lead to expanded and/or sustained practice changes without hospital pharmacy leader support.
2. This cross-sectional survey of hospital pharmacy leaders identified perceptions, resources and barriers to justifying clinical pharmacist positions.
3. This study highlights gaps in hospital pharmacy leader perceptions and knowledge, providing a foundation for developing evidence-based tools and targeted strategies to expand clinical pharmacy services, improve quality of care, and support clinician sustainability.

## Introduction

Clinical pharmacist services are shown to reduce medication errors, enhance medication optimization, and decrease adverse drug event rates, ultimately contributing to overall improvement of patient health outcomes.^1-6^ Although most hospitals in the U.S. provide some levels of clinical pharmacist coverage, the scope of services, staffing models, and workload expectations vary widely and are not standardized.^7,8^

Despite a substantial body of evidence demonstrating positive impact of clinical pharmacist services on patient outcomes, translating these pilot programs into sustained practice remains challenging and often requires additional clinical pharmacist positions (i.e., full time equivalents [FTEs]).^1-6,9-12^ Ongoing research continues to explore optimal clinical pharmacy practice models to improve patient outcomes and pharmacist well-being.^13^ These research efforts are only justified, however, if there is potential to implement the findings, which requires advocacy from hospital pharmacy leaders to add and/or reallocate FTEs to perform the clinical services evaluated.^11,12^ Drivers and barriers to expanding pharmacy services from the perspective of hospital pharmacy leaders and hospital executives (e.g., Chief Financial Officer, Chief Medical Officer, etc.) remain poorly understood.

The purposes of this study were to understand the perceptions of hospital pharmacy leaders regarding advocacy for expanded clinical pharmacy services and to identify factors deemed important for clinical pharmacist staffing (e.g., specific metrics or evidence). The findings from this study will inform future studies of clinical pharmacist services to promote evidence-based clinical pharmacy practice models that optimize medication use, enhance patient health outcomes, and improve clinician well-being.

## Methods

A national, prospective, cross-sectional survey was conducted to examine hospital pharmacy leader insights into supporting clinical pharmacist services. The survey was developed using the REDCap platform and distributed via email, web-based discussion board, or listserv to the American Society of Health-System Pharmacists (ASHP) Section of Pharmacy Practice Leaders, the American College of Clinical Pharmacy (ACCP) Critical Care Practice and Research Network (PRN) (with a request to forward to pharmacy administrators), and to the Georgia Society of Health-System Pharmacists (GSHP) Directors of Pharmacy. Individuals currently employed as hospital or health system leaders in the U.S. were eligible to participate. The survey was disseminated over a three-week period from September to October 2025 with a follow-up email sent one week prior to survey closure. The survey was approved by the University of Georgia Institutional Review Board (PROJECT00011879).

The survey consisted of 35 questions that were developed through an iterative, multi-step process of literature review, expert consultation, and cognitive pretesting. A literature search was conducted on PubMed to identify published manuscripts related to hospital pharmacy staffing, implementation of pharmacy practice model changes, expansion of clinical pharmacy services, expansion/implementation of services in other healthcare professions, and hospital administrator barriers and/or drivers to change staffing to explore conceptual domains and gaps not addressed in prior research. From these findings, initial concepts for the questionnaire were drafted and reviewed by five clinical pharmacist co-investigators to ensure content relevance and feasibility. Cognitive pretesting and pilot administration were tested by co-investigators and three pharmacy hospital leaders who were external to the project development to evaluate the clarity and validity of the questionnaire. Revisions were made until consensus was reached that the questions demonstrated adequate clarity and relevance to the intended purpose of the study.

Three primary domains were included in the survey: (1) perceptions on the climate related to clinical pharmacy at the respondent’s institution; (2) facilitators, barriers, and opportunities related to justification of clinical pharmacy services and FTEs; and (3) survey respondent and hospital/health system characteristics. The survey included a variety of item types, including Likert-scale, multiple-choice, and free-response questions. The definition of “clinical pharmacist” was provided on each page of the survey as follows: “For the purposes of this survey, a clinical pharmacist refers to a decentralized pharmacist who provides direct patient care as part of interprofessional healthcare teams. This includes pharmacists who round with medical teams in inpatient settings, manage medication therapy in outpatient clinics, and contribute to clinical decision-making. These individuals are often referred to as clinical specialists or advanced practice pharmacists.” For questions related to institutional characteristics, respondents representing a health system (rather than an individual hospital) were asked to answer based on their system’s largest hospital.

The primary outcome was identification of types of evidence most compelling to justify clinical pharmacist positions. Secondary outcomes included resources currently in use for decision-making and perceived barriers.

Likert-scale data were processed through IBP SPSS Statistics (Armonk, NY) and were reported using descriptive statistics. Quantitative responses were summarized using counts and percentages. Free-response items underwent thematic analysis, during which the responses were coded to identify recurring concepts and themes. Respondents with missing values for any question were excluded from analyses for that given question.

## Results

### Respondent and Institution Characteristics

The survey was answered by 123 healthcare administrators with 84 complete responses. Detailed data on baseline characteristics of survey respondents and hospitals/health systems is presented in **Table 1**. Over half of respondents identified as female, and 70% were between 30 and 49 years of age. Nearly all respondents had a Doctor of Pharmacy degree, and about one-third reported having non-pharmacy master’s degrees. The most common leadership roles of respondents were Director of Pharmacy Services and Clinical Manager/Supervisor.

**Table 1.**
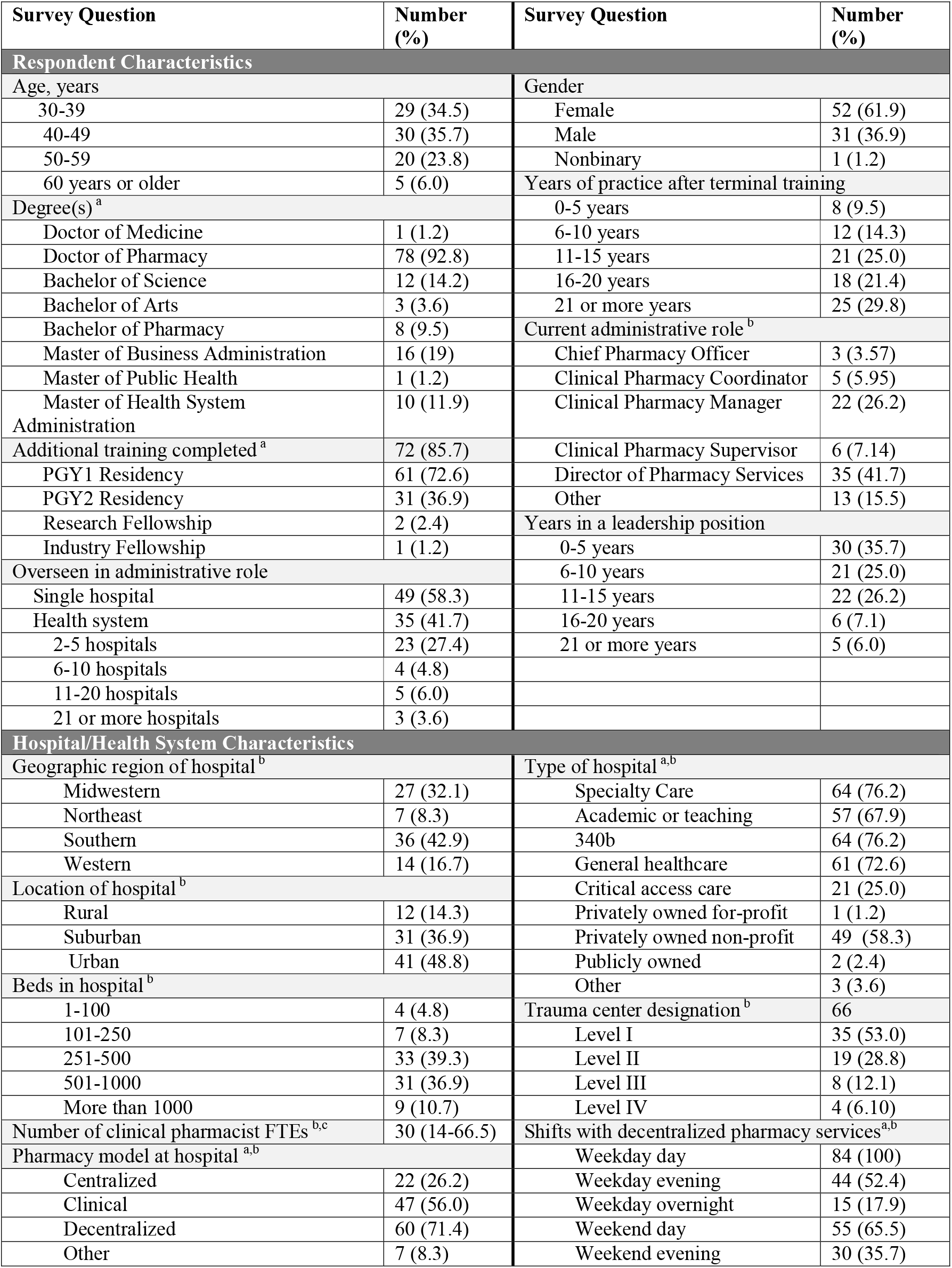
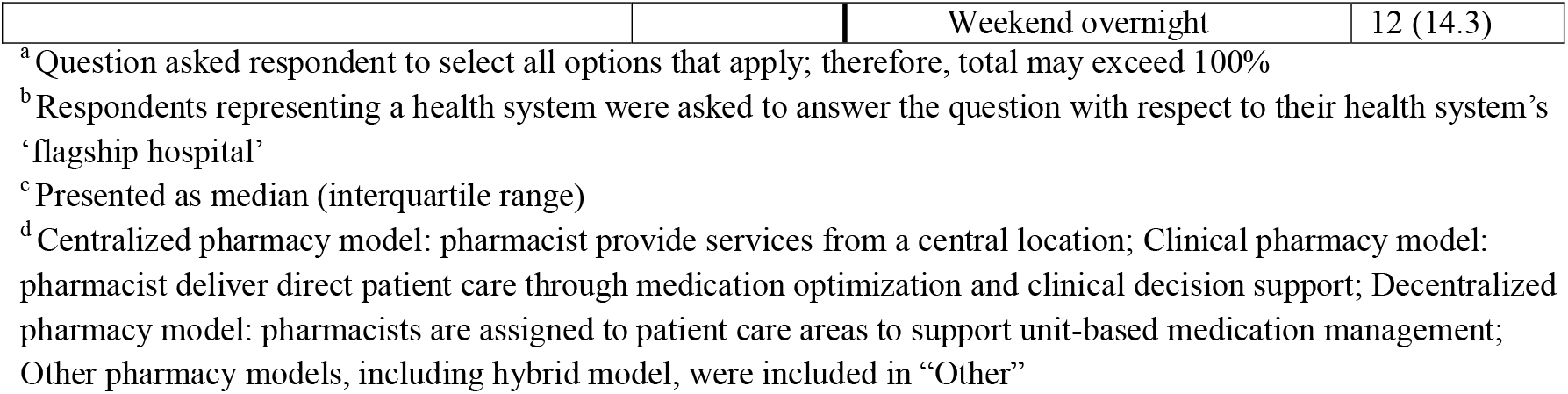
Respondent and Hospital/Health System Characteristics.

Respondent practice locations were distributed in all geographic regions of the U.S., with the South being most common. There was additionally diverse representation with regard to single hospitals versus health systems, hospital type (e.g., community, academic, government), and hospital size. Pharmacist staffing levels varied, with median of 30 FTEs. Decentralized pharmacy service models were common across institutions with weekday dayshift coverage for 100% of institutions using this model. Decentralized services were less common during other staffing times, with 65% on weekend dayshift, 52% on weekday evening shift, 36% on weekend evening shift, 18% on weekday night shift, and 14% on weekend night shift.

### Pharmacy Leader Perceptions & FTE justification

Pharmacy leaders reported mixed perceptions regarding adequacy and impression of clinical pharmacy services at their institutions (**Table 2**). When asked for their level of agreement with the statement “My hospital has enough clinical pharmacists for quality comprehensive medication management (CMM) for all patients” (scale of 1 [complete agreement] to 10 [complete disagreement]), the median rating was 6 (interquartile range [IQR] 4-8). There was high belief that additional clinical pharmacist FTEs would help decrease the workload for current employees (median score 3 [IQR 1-7]). Accordingly, 70% of pharmacy leaders reported requesting at least one new clinical pharmacy FTE within the previous 12 months, with a median of 3 (IQR 1-6) FTEs requested and 1 (IQR 1-3) approved. To justify these FTEs, the resources currently used by administrators include: benchmarking, productivity, or workload metric (65%); physician or other champions (63%); regulatory or accreditation requirements (62%); data indicating pharmacist impact on patient outcomes (62%), budget (61%), or hospital workflow (45%); quality indicators (e.g., Leapfrog results) (40%); improved patient access to care (35%); opportunity cost for the interdisciplinary team (25%); and employee satisfaction survey results (17%).

**Table 2.**
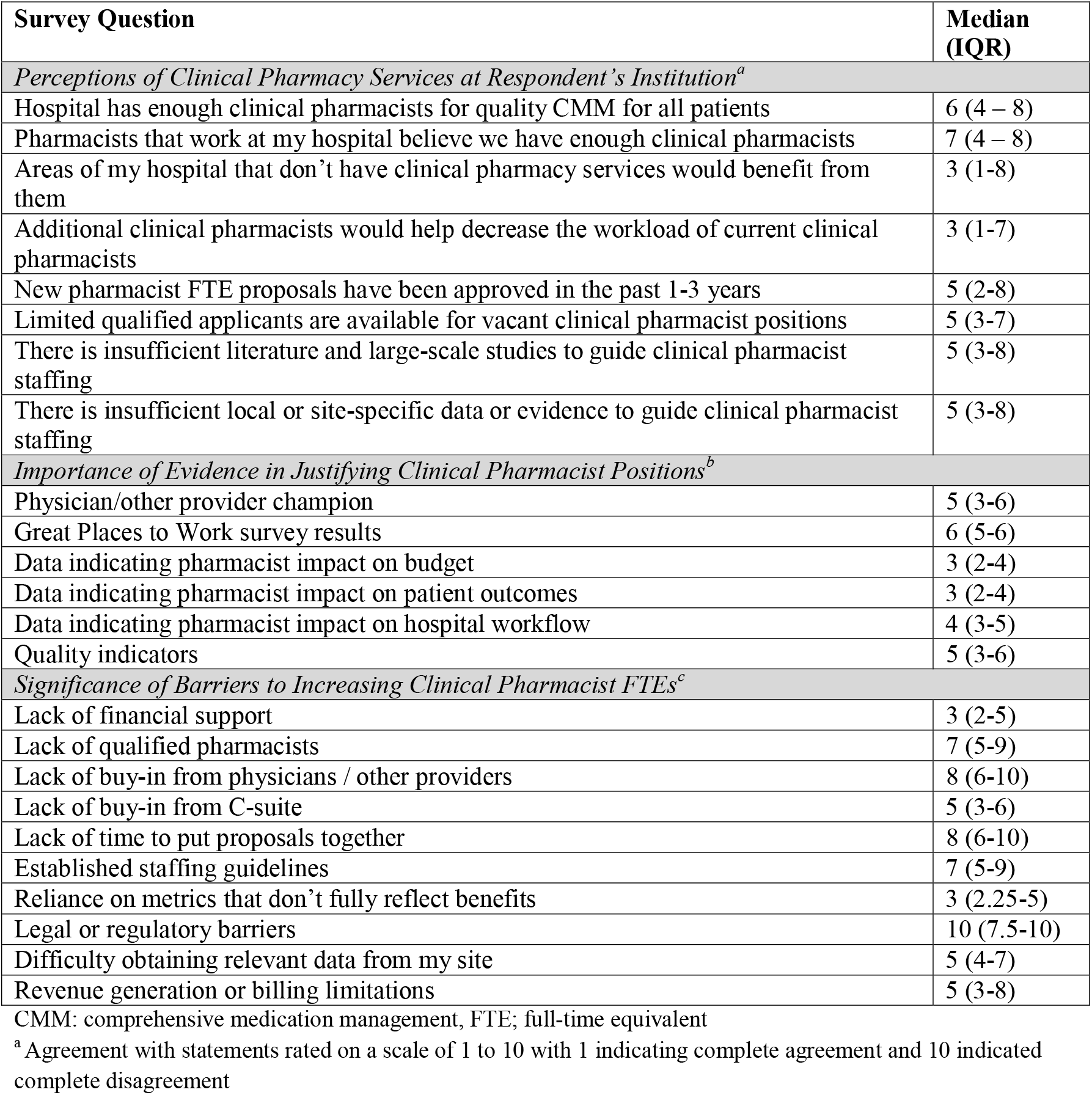
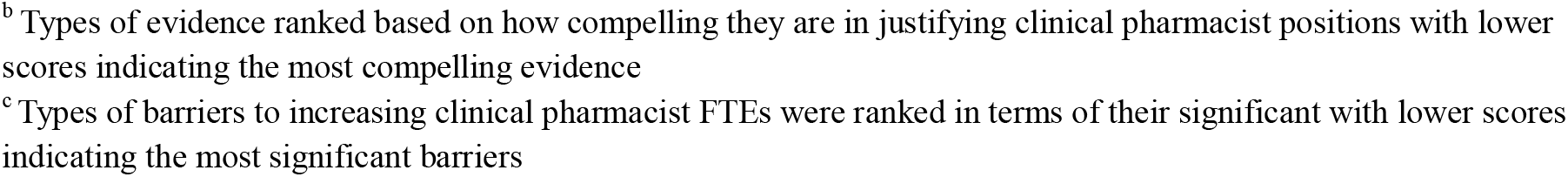
Pharmacy Administrator Perceptions on Clinical Pharmacy Services.

| Survey Question | Median (IQR) |
| --- | --- |
| *Perceptions of Clinical Pharmacy Services at Respondent's Institution<sup>a</sup>* |  |
| Hospital has enough clinical pharmacists for quality CMM for all patients | 6 (4 – 8) |
| Pharmacists that work at my hospital believe we have enough clinical pharmacists | 7 (4 – 8) |
| Areas of my hospital that don't have clinical pharmacy services would benefit from them | 3 (1-8) |
| Additional clinical pharmacists would help decrease the workload of current clinical pharmacists | 3 (1-7) |
| New pharmacist FTE proposals have been approved in the past 1-3 years | 5 (2-8) |
| Limited qualified applicants are available for vacant clinical pharmacist positions | 5 (3-7) |
| There is insufficient literature and large-scale studies to guide clinical pharmacist staffing | 5 (3-8) |
| There is insufficient local or site-specific data or evidence to guide clinical pharmacist staffing | 5 (3-8) |
| *Importance of Evidence in Justifying Clinical Pharmacist Positions<sup>b</sup>* |  |
| Physician/other provider champion | 5 (3-6) |
| Great Places to Work survey results | 6 (5-6) |
| Data indicating pharmacist impact on budget | 3 (2-4) |
| Data indicating pharmacist impact on patient outcomes | 3 (2-4) |
| Data indicating pharmacist impact on hospital workflow | 4 (3-5) |
| Quality indicators | 5 (3-6) |
| *Significance of Barriers to Increasing Clinical Pharmacist FTEs<sup>c</sup>* |  |
| Lack of financial support | 3 (2-5) |
| Lack of qualified pharmacists | 7 (5-9) |
| Lack of buy-in from physicians / other providers | 8 (6-10) |
| Lack of buy-in from C-suite | 5 (3-6) |
| Lack of time to put proposals together | 8 (6-10) |
| Established staffing guidelines | 7 (5-9) |
| Reliance on metrics that don't fully reflect benefits | 3 (2.25-5) |
| Legal or regulatory barriers | 10 (7.5-10) |
| Difficulty obtaining relevant data from my site | 5 (4-7) |
| Revenue generation or billing limitations | 5 (3-8) |

CMM: comprehensive medication management, FTE; full-time equivalent
<sup>a</sup> Agreement with statements rated on a scale of 1 to 10 with 1 indicating complete agreement and 10 indicated complete disagreement
<sup>b</sup> Types of evidence ranked based on how compelling they are in justifying clinical pharmacist positions with lower scores indicating the most compelling evidence
<sup>c</sup> Types of barriers to increasing clinical pharmacist FTEs were ranked in terms of their significant with lower scores indicating the most significant barriers

Respondents were asked to rank the types of evidence that would be most compelling to justify clinical pharmacist positions (even if the evidence isn’t currently available to them) and to rank the significance of different barriers in preventing clinical pharmacist FTEs from being approved (**Table 2**). Lower scores indicated a higher ranking (i.e., more compelling or a more significant barrier). The most compelling evidence identified was data indicating pharmacist impact on budget and patient outcomes. The most significant barriers identified were lack of financial support and reliance on metrics that don’t fully reflect benefits.

## Discussion

Health system pharmacy administrators from across the U.S. reported a strong need for additional clinical pharmacist positions and limited success in securing these FTEs, with only one-third of requested FTEs approved in the previous year. Administrators reported variability in the resources that they currently use to justify new FTEs with the most common being the pharmacist impact on patient outcomes and budget and other frequently used resources including performance and operational metrics, clinical impact, regulatory or accreditation requirements, and stakeholder (i.e., physician or other champion) influence. The variability observed in factors contributing to staffing decision-making suggests a reliance on subjective judgements, highlighting the need for a validated, data-driven framework to guide workforce determinations. This is further supported by the most significant barriers to obtaining approval for new clinical pharmacist FTEs, which were lack of financial support and the absence of reliable metrics to evaluate benefits of clinical pharmacists.

It is not surprising that the pharmacist impact on budget and lack of financial support were ranked amongst the most compelling evidence and most significant barriers to justifying clinical pharmacist FTEs, respectively. In current practice, both pharmacist salaries and drug costs are allocated to the pharmacy cost center. These siloed budgets can create a misalignment between costs and benefits. While clinical pharmacists contribute to improved patient outcome and provide substantial clinical, operational and financial benefits to hospitals, these benefits are not reflected within the pharmacy budget itself. Furthermore, justifying return on investment for inpatient clinical pharmacy services remains challenging because the addition of such services requires a significant financial commitment from institutions while not generating direct, billable revenue. Although clinical pharmacists undergo intensive and rigorous training to provide highly skilled, cognitively complex services as medication experts within the interprofessional team, these services are not currently reimbursed in the acute care setting. While progress has been made in some settings, such as ambulatory care,^14,15^ where pharmacists can bill for select services such as medication therapy management services or low level incident to provider billing, similar reimbursement models have not been widely adopted for acute care pharmacist services. Given the limitation of siloed budgets and the demonstrated value of clinical pharmacy services, alternative value-based funding approaches, such as service line-based funding or shared-saving models to reduce silo effects, should be explored to support the integration of clinical pharmacist FTEs into service expansion plans.^16,17^

Another significant barrier identified was the absence of reliable, meaningful metrics to evaluate the value of clinical pharmacists. Clinical pharmacy services are often assessed using outdated workload-based measures, such as the number of prescriptions verified or the number of documented consultations or interventions, which fail to capture the full scope of the services provided.^18^ Although these measures are easy to obtain, they primarily quantify activities rather than the impact of those services at a system or patient-specific level. From a broader perspective, pharmacist interventions reduce medication errors and prevent adverse drug events,^1,3,5,6,10^ thereby preventing costs associated with patient harm. In addition to value derived from direct patient care, indirect patient care contributions, including quality improvement, research, and professional services, should also be taken into account because they directly benefit the institutions and support sustainable, continuous advancement in care.^16^ These benefits are largely invisible when relying on traditional workload metrics. This gap emphasizes the need to develop more comprehensive, evidence-based metrics that reflect the full impact of clinical pharmacy services, including contributions to patient care quality, financial justification, and broader institutional benefit.

An interesting finding, consistent with observation from prior studies, was that decentralized clinical pharmacy practice has not yet become a standard of practice across US healthcare systems.^19^ Although most contemporary literature supports decentralized pharmacy practice models as superior to centralized models in terms of clinical impact, operational efficiency and patient outcomes,^13,19^ only 71% of respondents reported using a decentralized pharmacy model. The availability of decentralized clinical pharmacists during evening and overnight shifts and on weekends was far lower than weekday day shifts, despite evidence suggesting the positive impact of pharmacists on both patient care and cost-savings throughout all days of the week and staffing shifts.^16,20^

These findings provide fundamental principles for future research to develop a standardized framework justifying clinical pharmacist FTEs. Future studies should move beyond descriptive workload measures and focus on validating metrics that capture the full value of clinical pharmacists to both patient care and health system operations. Those metrics should include financial benefits or healthcare utilization from improved patient outcomes, along with measures of efficiency and resource utilization. Other outcomes that are less tangible but highly relevant to administrators, including improvements in patient satisfaction, enhancement of medication safety culture, and contribution to more reliable and progressive care environment, should also be considered. Ultimately, this approach would support data-driven decision-making and strengthen sustained investment in clinical pharmacy services.

This study represents the first national survey designed to improve clinical pharmacy staffing by evaluating hospital pharmacy leader perceptions, their rationale for FTE justification, and identifying barriers to expanding clinical pharmacy services. This survey captured responses from administrators across all regions in the United States, enhancing the external validity of the findings and providing a valuable foundation for future research to standardized approaches to clinical pharmacist staffing.

Several limitations should be noted. The sample size was relatively small, which may limit the representativeness of the results. Although differences in staffing needs and decision-making processes likely exist across various institutional settings (e.g., academic medical center versus critical access hospital), the limited number of responses prevented meaningful sub-analyses by hospital type. Additionally, the listservs used for survey distribution were selected based on their availability to study investigators and their potential to reach pharmacy leaders. A larger and more diverse platform for survey dissemination may have increased sample size and generalizability, and the use of one listserv specific to the state of Georgia may have impacted the larger representation of respondents from the Southeast. Furthermore, participation was voluntary, which may introduce response bias if administrators with strong opinions about clinical pharmacy staffing were more motivated to participate. Of those who responded, some hospital pharmacy leaders might choose not to request additional FTEs due to anticipated denial, leading to an overestimation of approval rate for clinical pharmacist FTEs. The approval rate of requests for FTEs of other professions in the hospital would be useful data to contextualize our findings, but is not available in the literature. Despite these limitations, this survey provides an important snapshot of pharmacy administrator perceptions. Future research with larger and more diverse samples is needed to validate and expand upon the findings of this study.

## Conclusion

This study offers insight into hospital administrator perspectives regarding the resources and evidence required to support clinical pharmacy practice changes and justification of new FTEs. Although patient outcome and cost-related measures strongly influence administrative decision-making, existing metrics do not fully capture the comprehensive value of clinical pharmacy services, indicating a need for novel metrics and a framework to support implementation of optimal clinical pharmacy practice models. By highlighting areas of potential gaps in meaningful evidence, this study provides guidance for future research designed to advance clinical pharmacy services, improve quality of care, and support clinician sustainability.

## Data Availability

All data produced in the present study are available upon reasonable request to the authors

## Notes

**Conflicts of Interest:** The authors have no conflicts of interest.

### Competing Interest Statement

The authors have declared no competing interest.

### Funding Statement

Funding through the Agency for Healthcare Research and Quality for Drs. Sikora and Smith was provided through R21HS028485 and R01HS029009.

### Author Declarations

Institutional Review Board of the University of Georgia gave ethical approval for this work (PROJECT00011879).

